# Features and outcomes of secondary sepsis and urinary tract infections in COVID-19 patients treated with stem cell nebulization

**DOI:** 10.1101/2020.12.05.20244483

**Authors:** Gina M. Torres Zambrano, Rene Antonio Rivero, Carlos A. Villegas Valverde, Yendry Ventura Carmenate

**Author notes:** Mailing address: Street #31, Area Muroor Road, Building # C43, Al Murjan Tower, Apartment 107, Abu Dhabi, UAE. Abu Dhabi Stem Cells Center (ADSCC) Ringgold ID: 580443.

## Abstract

**Background:** COVID-19 is the defining global crisis of our time. Secondary complication such as urinary tract infections and sepsis, worsen the already established problem, creating a new challenge.

**Objective:** To characterize the features and outcomes in COVID-19 patients with sepsis and urinary tract infection.

**Methods:** An observational and analytical study was conducted within the framework of the SENTAD COVID clinical trial at the Abu Dhabi Stem Cells Center, were the patients received a nebulization therapy with the use of autologous stem cells (group A). Those patients were compared with a not stem cells treated control arm (group B), and both received the UAE COVID 19 standard management. An analysis of the culture samples, antimicrobial agents and the efficacy of the therapy on patient’s outcomes was done.

**Results:** A significant difference between the groups was found in the UTI incidence (p=*0.0206). Patients in group A showed a lower tendency to sepsis in comparison with group B (7% vs 21%), HR=0.35, (95% Confidence Interval: 0.13 – 0.91), p=0.0175. It was calculated a NNT=7.3. *Candida albicans* was the most frequently agent causing sepsis and UTI. The massive use of broad-spectrum antimicrobials was striking.

**Conclusions:** We found a protective factor of stem cells against secondary infection in COVID 19 cases, in terms of sepsis and UTI. The suggested immunomodulatory effect of stem cells offers a therapeutic strategy to manage the disease and avoid several complications. Antimicrobial agents can lead to increased opportunistic infections, so a rational approach to these treatments must be considered.

## Introduction

The corona virus disease (COVID-19) is the defining global crisis of our time and the greatest threat this world has faced in a long time. Many countries, including the UAE are taking every precautionary measure to slow and control the spread.^1^

Taking into account that the sepsis and septic shock contribute to the cytokine storm syndrome causing a high mortality rate (60–90%)^2^ and also generate an immune-mediated kidney damage,^2,3^ it is crucial to prevent this event. Studies showed that there was a sharp increase in Methicillin-resistant *Staphylococcus aureus* (MRSA), *Pseudomonas* and *Candida* species infectious rate among the intensive care unit (ICU) COVID-19 patients, indicating that secondary infection affect the prognosis and subsequent treatment of these patients.^4^

Given the immunomodulatory potential of the stem cells in sepsis and clinical evolution of chronic conditions with the stimulation by humoral factors, strategies such as stem cell-based therapy are being proposed to regulate inflammation, prevent or mitigate this cytokine storm through their immunomodulatory capacity.^5^

The Abu Dhabi Stem Cell Centre (ADSCC) research team developed a treatment for COVID-19 (“SENTAD COVID Study” clinical trial) with the use of an autologous stem cells cocktail, called Peripheral Blood Non-Hematopoietic Enriched Stem Cells (PBNHESC), that has shown promising results.^6^ Previously, we studied the frequency of acute renal injury (AKI) in patients with COVID-19 and their relation with clinical outcomes within the framework this clinical trial, finding it in about a third of critically ill patients. In addition, those who received the treatment showed a better tendency to improve in terms of hospital stay and biomarker evolution (amelioration of linphopenia,^7–9^ neutrophil-lymphocyte ratio^10–13^ and C-reactive protein (CRP)) than those of the control group.

Scientist and researchers all over the world are racing to find treatment because not only is COVID-19 a major health crisis, but it also has the potential to create devastating social, economic and political catastrophes to every country it touches. In this article we analysed the incidence of sepsis and urinary tract infection (UTI) and related pathogens, in comparison with the rest of the germs that cause sepsis in the two groups of patients of our study, after receiving the treatment or being recruited as a control.

## Objective

To characterize the features and outcomes in COVID-19 patients with sepsis and urinary tract infection after intervention, within the framework of the ADSCC SENTAD COVID clinical trial at the Abu Dhabi Emirate, during the month of April of 2020.

## Methods

Within the framework of the SENTAD COVID Study clinical trial,^6^ we performed a multi-center, prospective and analytical study to figure out the sepsis and UTI incidence among the patients after the intervention. Four different hospitals from the Emirate of Abu Dhabi participated: Sheikh Khalifa Medical City, Al Rahba Hospital, Al Mafraq Hospital and Al Ain Hospital. From the main study, a total of 139 patients were divided in two groups for the comparison:

1. Group A (Experimental arm) COVID 19 standard care plus nebulization with PBNHESC (n=69)
2. Group B (No intervention arm) COVID 19 standard care (n=70).

### Both groups fulfil the inclusion criteria

Aged ≥ 18 years, laboratory confirmation of COVID-19, interstitial lung change judged by computed tomography, hospitalized and symptomatic patients, referring one or more symptoms, ability to comply with test requirements and blood collection and agrees to participate in the study.

### The exclusion criteria were

Patients with diagnosis of any kind of shock, organ transplants in the past 3 months, patients receiving immunosuppressive therapy, diagnostic of Hepatitis B Virus (HBV) infection or Human Immunodeficiency Virus (HIV) infection or Acquired Immunodeficiency Syndrome (AIDS), current diagnosis of cancer or history of malignancies in the past 5 years, pregnant or lactating women, patients who had participated in other clinical trials in the past 3 months, inability to comply with test requirements and blood collection or inability to provide informed consent.

In order to determine the clinical severity, the ordinal scale created by the WHO committee for COVID -19, that measures the illness severity overtime, was used:^14^ (Table 1)

**Table 1.** SENTADO-COVID Study seven-category ordinal scale for clinical involvement.

| Category | Score |
| --- | --- |
| No limitation of activities, discharged from hospital | 1 |
| Limitation of activities. | 2 |
| Hospitalized, no oxygen therapy. | 3 |
| Oxygen by mask or nasal prongs. | 4 |
| Non-invasive ventilation or high-flow oxygen | 5 |
| Intubation and mechanical ventilation. | 6 |
| Mechanical Ventilation + additional organ support: ECMO, CRRT, vasopressors. | 7 |
| Death | 8 |

A registration of gender, age, BMI, and comorbidities, vital signs, and biochemical studies performed within 24 h of inclusion in the trial was done. Both groups received COVID 19 standard treatment defined as “UAE National Guidelines for Clinical Management and Treatment of COVID 19, v.2.0” as per the Department of Health (DOH)^1^. A stem cell nebulization with autologous PBNHESC-C (cocktail rich in very small embryonic-like stem cells (VSELs) and platelets rich plasma derived growth factors, commercially named *UAECell 19*), was given by two nebulization of 10cc, in two consecutive days (at least 22 hours between each one) for patients at group A, obtained through phlebotomy of 300cc and processed at the ADSCC laboratory, and cells were characterized by flow cytometry and inverted automated fluorescent microscopy.

Follow up of both groups were done until the discharge. Secondary infection was diagnosed when the patients had clinical symptoms with a positive culture (blood, sputum, others) and UTI with a positive urine culture. Consequently, the reports of the sputum, blood, specific seeking of Methicillin-resistant *Staphylococcus aureus* (MRSA), skin/wound and urine cultures were collected, as well as the antimicrobial treatment given.

### Statistical analysis

a non-normal distribution of the variables was found, so non-parametric statistical methods were used. A proportions comparison test (Chi-Square) for the body mass index (BMI) categories^15^ and comorbidities, and *U*-Mann-Whitney for the dates intervals. The Hazard Ratio (HR), 95% Confidence Interval, Number Needed to Treat (NNT) was calculated for the sepsis complication. A significance level of p <0.05 was prefixed.

### Ethics

The study was approved by the Emirates Institutional Review Board for COVID-19 Research (Ref. ID: DOH/CVDC/2020/1172). Study participants provided written informed consent per the Helsinki Declaration.^16^ The consent document described the importance of participation and explained the study’s characteristics and possible risks and benefits. All data were kept confidential and participant identity was delinked. The selection of diagnostic tools followed the ethical principles of maximum benefit and non-maleficence. This work was supported by ADSCC. This research had not received any other specific grant from any funding agency in the public, commercial or not-for-profit sectors. Award/Grant number is not applicable. None of the contributing authors have any conflicts of interest. For extra data.

## Results

From the 139 patients, there were 129 males (93%) and 10 females (7%), 65 males (94%) from the group A and 64 (91%) from the group B, and 4 females (6%) vs. 6 (9%) correspondingly. The ages of group A ranged from 27 to 71 years old (mean 45.9 years old), and from 26 to 73 years old in group B (mean 44.31 years old), with no significant differences between the two groups (p=0.3677). The distribution of the BMI was between 17.06 to 45.44 (mean 27.61) in the group in the group A, and between 16.95 to 47.75 B (mean 27.93), although there were more patients with overweight in group A. (Table 2)

**Table 2.** Clinics characteristics of the patients.

|  | Group A |  | Group B |  | p* |
| --- | --- | --- | --- | --- | --- |
|  | n | % | n | % |  |
| BMI: |  |  |  |  |  |
| Overweight (25-29) | 35 | 51% | 19 | 27% | **0.0038** |
| Mild obesity (30-34) | 11 | 16% | 13 | 19% | 0.6429 |
| Moderate obesity (35-40) | 0 | 0% | 3 | 4% | 0.0945 |
| Morbid obesity (>40) | 3 | 4% | 4 | 6% | 0.5901 |
| Comorbidities |  |  |  |  |  |
| Smoker | 6 | 9% | 1 | 1% | **0.0306** |
| DM | 18 | 26% | 13 | 19% | 0.3246 |
| HTN | 18 | 26% | 19 | 28% | 0.7913 |
| DL | 5 | 7% | 7 | 10% | 0.5277 |
| Cardiac diseases | 1 | 1% | 2 | 3% | 0.4023 |
| Respiratory diseases | 4 | 6% | 3 | 4% | 0.5896 |
| Score |  |  |  |  |  |
| 3 | 36 | 52% | 40 | 59% | 0.4080 |
| 4 | 13 | 19% | 6 | 7% | 0.0252 |
| 5 | 3 | 4% | 6 | 10% | 0.0960 |
| 6 | 2 | 3% | 3 | 4% | 0.7494 |
| 7 | 15 | 22% | 15 | 21% | 0.8863 |

BMI: Body Mass Index. DM: Diabetes Mellitus. HTN: Hypertension. DL: Dyslipidemia. \*: proportions comparison test.

Diabetes Mellitus and hypertension were the most commons comorbidities in both groups. No differences were found between the groups regarding those variables. There were more smokers in the group A (1% vs 9%, *p=0.0306). Likewise, there were no differences in the scores between the groups. Table 2 is a summary of the data.

The median hospital stay after receiving the treatment in the study for group A was 5 days, and 6 days in group B after being included, but the range for group B was from 2 to 125 days, in compared with 1 to 43 days in group A (p= 0.2924). Apropos the UTI incidence, a significant difference between the groups was found (p=0.0206). Patients in group A showed a lower tendency to sepsis in comparison with group B (7% vs 21%), HR=0.35, (95% Confidence Interval: 0.13 – 0.91), p=0.0175. It was calculated a NNT=7.3. In group B there were 6 deaths while in the group A there were only 4. (Table 3)

**Table 3.**
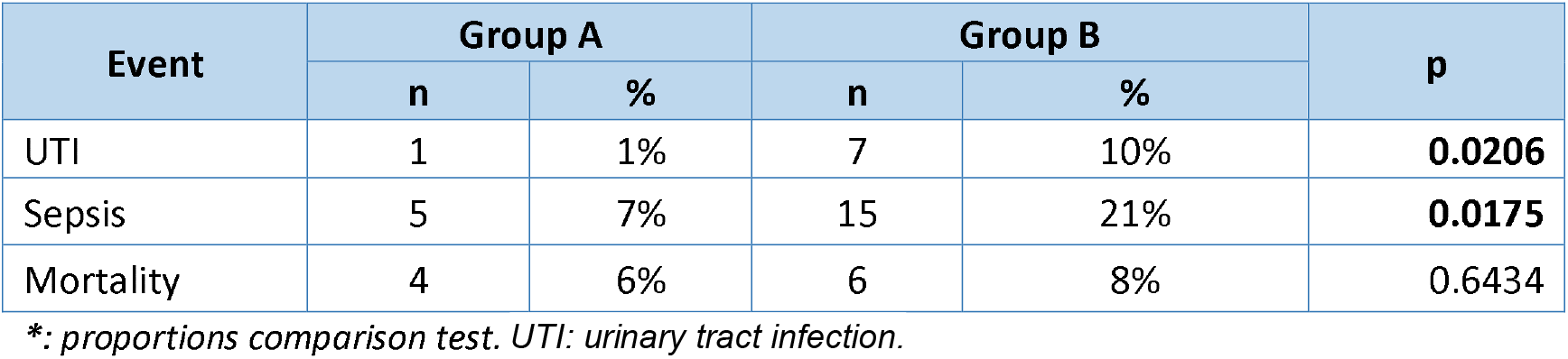
Clinical outcome characterizations.

| Event | Group A |  | Group B |  | p |
| --- | --- | --- | --- | --- | --- |
|  | n | % | n | % |  |
| UTI | 1 | 1% | 7 | 10% | **0.0206** |
| Sepsis | 5 | 7% | 15 | 21% | **0.0175** |
| Mortality | 4 | 6% | 6 | 8% | 0.6434 |

\*: proportions comparison test. UTI: urinary tract infection.

Cultures were obtained from all the critically ill patients, 23 from group B and 20 from group A, 41 samples from blood (23 vs. 18), 23 from urine (22 vs. 1), 31 MRSA (22 vs. 9), 29 from sputum (15 vs. 14) and 2 from wound/skin (2 vs. 0). In total 39 patient’s samples (28%) identified at least one pathogen. From group B, 27 (38%) samples were positive: 9 from blood (39%), 8 from urine (36%), 6 from sputum (40%), 1 MRSA (5%) and 3 from wound/skin (100%). From group A, 12 samples were positive (17%), 5 from blood (28%), 1 from urine (5%), 6 from sputum (43%).

*Candida albicans* caused 21% of the sepsis in both groups (7 from group B vs 2 from group A), *Staphylococcus aureus* 19% (5 vs 3 cases), *Pseudomonas aeruginosa* 16% (3 vs 4 cases), *Streptococcus pneumonia* 5% (1 vs 1 case), *Klebsiella pneumoniae* 9% (4 vs 0 cases), *Escherichia coli* 9% (4 vs 0 cases) and *Enterobacter aerogenes* 2% (1 vs 0 cases). From the group A, 6 patients with UTI were found to have other positive cultures, 3 from blood, 2 from sputum and 1 had MRSA. The patient from group A who had UTI had sepsis caused by *Candida albicans*. Of the deceased patients, only one from group B had a UTI (14%) caused by *Klebsiella pneumoniae*, none from group A. (Figure 1)

**Figure 1.**
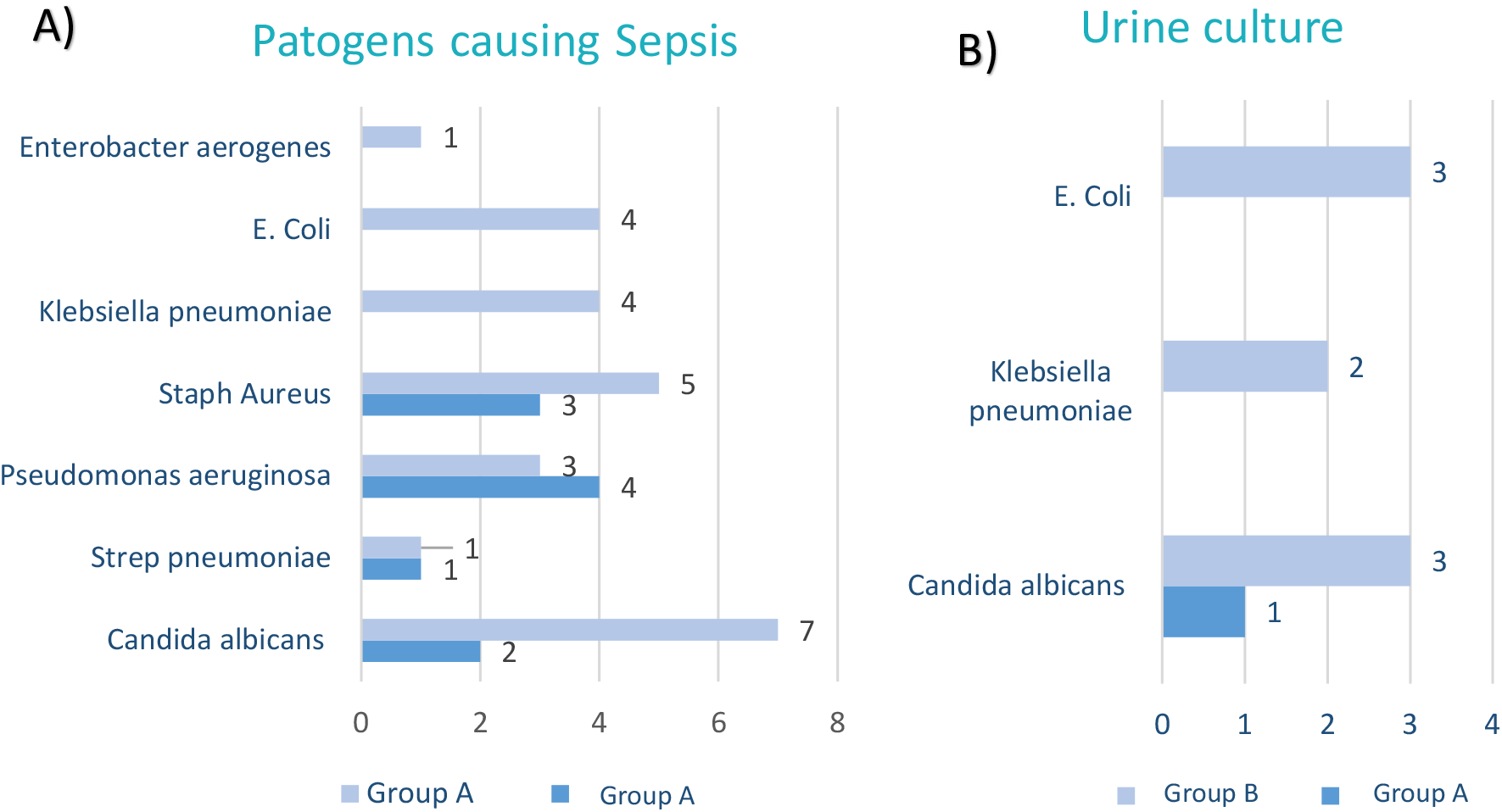
Cultures reports: A) Number of cases with pathogens found in different cultures Y B) Number of cases with pathogens grown in urine cultures.

In 11 cases more than one pathogen grew, 9 from group B and 2 from group A, where *Candida albicans* was the most frequently found concomitant pathogen along with other germs (5 vs.2), followed by *Staphylococcus aureus* (3 vs. 1), *Klebsiella pneumonia* (3 vs. 0) and *Pseudomonas aeruginosa* (2 vs. 1). (Table 4)

**Table 4.** Multiple pathogens infection distribution.

| Pathogens | Group A | Group B |
| --- | --- | --- |
| *Candida albicans* + *Pseudomonas aeruginosa* | 1 | 1 |
| *Candida albicans* + *Staphylococcus aureus* | 1 | 1 |
| *Candida albicans* + *Klebsiella pneumoniae* |  | 1 |
| *Candida albicans* + *Streptococcus pneumoniae* |  | 1 |
| *Candida albicans* + *Enterobacter aerogenes* |  | 1 |
| *Klebsiella pneumoniae* + *Acinetobacter baumannii* |  | 1 |
| *Streptococcus pneumoniae* + *Klebsiella pneumoniae* + *Escherichia coli* |  | 1 |
| *Staphylococcus aureus* + *Escherichia coli* |  | 1 |
| *Pseudomonas aeruginosa* + *Staphylococcus aureus* + *Enterobacter aerogenes* |  | 1 |
| Total | 2 | 9 |

Regarding the use of antibiotics, 60 pacients received this therapy, 30 from each group. Piperacillin/tazobactam was the most commonly used in both groups, followed by Meropenem, Vancomicin and Linezolid. (Figure 2). It was marked the use of several antimicrobial agens in 42 patients (30%), 17 from group A (25%) and 25 patients from group B (35%), from those, 11 received 2 combination of antibiotics (8%, 9 from group A and 7 from group B), 15 received 3 (11%, 11 vs. 4), 6 received 4 (4%, 4 vs. 2), 3 received 5 (2%, 3 vs. 0) and 2 received 6 (1%, 2 vs. 0).

**Figure 1.**
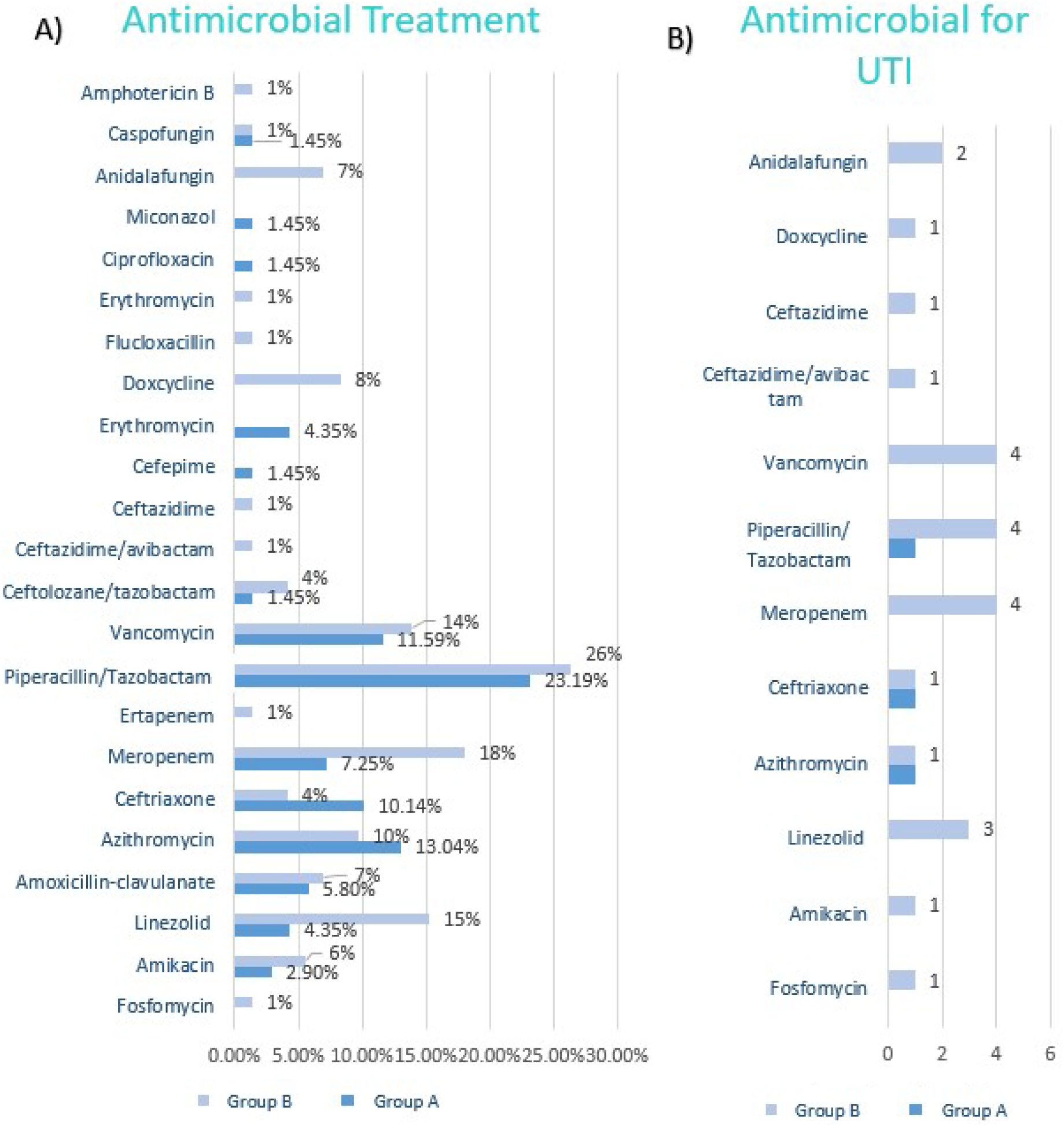
Antimicrobial treatment. A) Treatment for from both groups. B) Treatment for patients with UTI.

On the subject of the antifungical treatment, 9 patient received treatment, 2 from group A and 7 from group B. From the second group, 1 patient received 2 antifungical (this patient received also treatment with linezolid and meropenem). The most frequently used in group B was anidalafungin and caspofungin was in group A. (Figure 2)

## Discussion

The data analyzed from this study showed a higher incidence of COVID-19 among men, especially among critically ill patients (no women with high scores were found during the study period). Other authors confirmed male predominance in the incidence of this disease compared to female, it has also been found that patients with diabetes, hypertension, coronary heart disease, chronic obstructive pulmonary disease, cerebrovascular disease, and kidney disease exhibit worse clinical outcomes when are infected with COVID-19^17,18^. Similarly, the patients included in the study were found to have this comorbidities as risk factors forsepsis. ^19^ Despite the fact that among the treated patients there were more smokers and more overweight cases, it was found that infectious complications were lower than those of the control group.

Bacterial and fungal secondary infection rate in hospitalized adults with COVID-19 is poorly understood, and are associated with intensive care unit admission and higher risk of invasive procedures^4^. As a result, secondary infection might be a key element that leads to severe disease and mortality. In our study, there was high culture sampling rate among he critically ill patients in both arms, with an important role of the *Candida albicans* causing sepsis (21% of cultures) and was the most common germ in association with other bacterial infections, but treated patients were less prone to this event than controls, with a statically significant difference, and it was found that around 7 patients are required to be treated to have this protective effect with the stem cell nebulization (NNT=7.3). Surprisingly, despite the fact that all septic patients had a urinary catheter, the rate of UTI was not strikingly high in group A compared with group B. An attenuation of the bacterial sepsis stem cell mediated had been described, via several mechanisms such as improving the phagocytic ability, secreting anti-microbial peptides^5,20^, and increasing bacterial clearance.^5,21^

The COVID-19 sepsis can be associated with an immunosuppresive effect of the SARS-Cov-2 and for instance, as shownin this study *C Albicans,as well as Pseudomonas*, are more frequenty found as oportunistic patogen associted with a depressed immune response. Taking into count that a large proportion of patients had received therapy with one or more broadspectrum antibacterials at the time of culture, in some cases up to 6 antibiotics, the reason of the fungal sepsis could be related to these managements. Other authors also found evidenced a strong increase in opportunistic infections rate among SARS ICU patients, due to MRSA, *Pseudomonas*, and *Candida* species.^4,22^ There is being described the biofilm formation as a resistant mediator, especially in those with MRSA positive culture, which is a survival mechanism, since it constitutes a reservoir of bacteria with multi-resistance.23,24 The antibacterial therapy, if indicated, should be prescribed in line with local guidelines and reviewed with clinical response at 48 to 72 hours. If no evidence of bacterial coinfection is found, then stopping antibacterial therapy should be considered.^25^ It is well known that the prolonged use of antimicrobials and their inappropriate use, contribute to the development of major consequences, and we are facing a lack of data about antimicrobial use during this pandemic worldwide in just a few months, then the emergence of new resistance to this treatmets should be expected, without counting other adverse effects of these therapies for kidney and liver function for example, with unimaginable implications for human and animal health and the environment.^26^

Another reason of these secondary infections must be related with the lymphopenic status of the critically ill COVID-19 patients already found in the main study^6^. There is a sustained and substantial reduction of the peripheral lymphocyte counts, especially CD4 T and CD8 T cells, as the representative of immune suppression stage after the cytokine storm activation phase increasing the risk of developing secondary infection^4,27^.

Even though the median hospital stay after the intervention was not significant between the groups, the range is much shorter in group A patients than in group B, given the homogeneity of the treated group due to the therapy, and the statistical dispersion of the data of the control group, giving another hypothesis of the lower incidence of sepsis and UTI in group A, since the shorter hospital stay is a protective factor against nosocomial infections^28^.

Moreover, due to the autologous origin of the therapy, there are no immunologic reactions, with rare adverse effects more linked with the collection of the blood (fainting and dizziness may occur).

There are some limitations to the current study. It was carried in 4 different hospitals, each one with different criteria for taking cultures, as well the selection of the initial empiric antimicrobials treatment, can cause multiple germ resistant and negativization of some of the samples. It is our responsability for implementing and developing actions to control or stop the disease, but also change dangerous practices, through appropriate and consensual use of antimicrobials.

## Conclusion

In this study, we found a protective factor of stem cells against secondary infection in COVID 19 cases, in terms of sepsis and UTI. Patients in the treated group showed a lower tendency to develop these events compared to the control group. The suggested immunomodulatory effect of stem cells offers a therapeutic strategy to manage the disease and avoid several complications, becoming a crucial adjuvant tool for healing and achieving early recovery in severe COVID-19 infections. It is well known that during this disease, patients had received several antimicrobial agents that can lead to increased fungal and opportunistic infections, so a rational approach to these treatments should be considered.

## Take Home Message

There is a significant frequency of sepsis and UTI as complications in COVID-19, related to the antimicrobial use, length of hospital stays and invasive procedures. Novel stem cell nebulization therapy may improve the clinical evolution and frequency of this complications.

## Data Availability

Extra data is available by emailing.

## Acknowledgments

Acknowledgment to the government of the United Arab Emirates and especially to the Abu Dhabi Health Services Company SEHA for the contribution and support to this study.

## Notes

### Competing Interest Statement

The authors have declared no competing interest.

### Clinical Trial

NCT04473170

### Clinical Protocols

https://clinicaltrials.gov/ct2/show/study/NCT04473170?term=stem+cell&cond=covid&cntry=AE&draw=2&rank=1.

### Author Declarations

The study was approved by the Emirates Institutional Review Board for COVID-19 Research (Ref. ID: DOH/CVDC/2020/1172).

